# COVID-19 booster vaccination during pregnancy enhances maternal binding and neutralizing antibody responses and transplacental antibody transfer to the newborn (DMID 21-0004)

**DOI:** 10.1101/2022.06.13.22276354

**Authors:** Flor M. Munoz, Christine M. Posavad, Barbra A. Richardson, Martina L. Badell, Katherine Bunge, Mark J. Mulligan, Lalitha Parameswaran, Clifton Kelly, Courtney Olsen-Chen, Richard M. Novak, Rebecca C. Brady, Marcela Pasetti, Emily DeFranco, Jeffrey S. Gerber, Mallory Shriver, Mehul S. Suthar, Kathryn Moore, Rhea Coler, Bryan Berube, So Hee Kim, Jeanna M. Piper, Ashley Miller, Cristina Cardemil, Kathleen M. Neuzil, Richard Beigi, DMID Study Group

**Author notes:** Corresponding Author: Flor M. Munoz, MD, Baylor College of Medicine, Texas Children’s Hospital, 1102 Bates Ave, Suite 1150, Houston, TX 77030. DMID 21-0004 Study Group.

## Abstract

**Importance:** COVID-19 vaccination is recommended during pregnancy for the protection of the mother. Little is known about the immune response to booster vaccinations during pregnancy.

**Objective:** To measure immune responses to COVID-19 primary and booster mRNA vaccination during pregnancy and transplacental antibody transfer to the newborn.

**Design:** Prospective cohort study of pregnant participants enrolled from July 2021 to January 2022, with follow up through and up to 12 months after delivery.

**Setting:** Multicenter study conducted at 9 academic sites.

**Participants:** Pregnant participants who received COVID-19 vaccination during pregnancy and their newborns.

**Exposure(s):** Primary or booster COVID-19 mRNA vaccination during pregnancy.

**Main Outcome(s) and Measure(s):** SARS-CoV-2 binding and neutralizing antibody (nAb) titers after primary or booster COVID-19 mRNA vaccination during pregnancy and antibody transfer to the newborn. Immune responses were compared between primary and booster vaccine recipients in maternal sera at delivery and in cord blood, after adjusting for days since last vaccination.

**Results:** In this interim analysis, 167 participants received a primary 2-dose series and 73 received a booster dose of mRNA vaccine during pregnancy. Booster vaccination resulted in significantly higher binding and nAb titers, including to the Omicron BA.1 variant, in maternal serum at delivery and cord blood compared to a primary 2-dose series (range 0.55 to 0.88 log_10_ higher, p<0.0001 for all comparisons). Although levels were significantly lower than to the prototypical D614G variant, nAb to Omicron were present at delivery in 9% (GMT ID50 12.7) of Pfizer and 22% (GMT ID50 14.7) of Moderna recipients, and in 73% (GMT ID50 60.2) of boosted participants (p<0.0001). Transplacental antibody transfer was efficient regardless of vaccination regimen (median transfer ratio range: 1.55-1.77 for binding IgG and 1.00-1.78 for nAb).

**Conclusions and Relevance:** COVID-19 mRNA vaccination during pregnancy elicited robust immune responses in mothers and efficient transplacental antibody transfer to the newborn. A booster dose during pregnancy significantly increased maternal and cord blood antibody levels, including against Omicron.

Findings support continued use of COVID-19 vaccines during pregnancy, including booster doses.

**Trial Registration:** clinical trials.gov; Registration Number: NCT05031468; https://clinicaltrials.gov/ct2/show/NCT05031468

**Key Points:** *Question:* What is the immune response after COVID-19 booster vaccination during pregnancy and how does receipt of a booster dose impact transplacental antibody transfer to the newborn?

*Findings:* Receipt of COVID-19 mRNA vaccines during pregnancy elicited robust binding and neutralizing antibody responses in the mother and in the newborn. Booster vaccination during pregnancy elicited significantly higher antibody levels in mothers at delivery and cord blood than 2-dose vaccination, including against the Omicron BA.1 variant.

*Meaning:* COVID-19 vaccines, especially booster doses, should continue to be strongly recommended during pregnancy.

## Introduction

Pregnant individuals are at increased risk of severe disease after SARS-CoV-2 infection.^1,2^ Infants younger than 6 months of age who become infected with SARS-CoV-2 are at increased risk of hospitalization.^3^ COVID-19 vaccination during pregnancy is critical to mitigate the burden of disease for mothers and their infants.^4^ In October 2021, pregnant individuals became eligible for booster vaccinations, yet the response to a booster dose has not been well characterized.^5^

In this prospective cohort study, we measured the antibody response to COVID-19 mRNA vaccines in pregnant participants and antibody levels in cord blood. Here, we report an interim analysis of the effect of booster vaccination in a subset of mothers and on transplacental antibody levels in the newborn.

## Methods

This U.S. (United States)-based multicenter study enrolled pregnant participants with and without medical comorbidities from July 6, 2021 to January 31, 2022. Eligible participants received a primary 2-dose series of Pfizer-BioNTech (Pfizer) or Moderna mRNA-1273 (Moderna) vaccine, or a booster dose of either vaccine, at any time during pregnancy as per current recommendations. Sera for antibody assays were derived from maternal blood collected pre- and post-vaccination (from 2 weeks post-vaccination to delivery), and maternal and cord blood collected at delivery. Detailed protocol and study procedures are described elsewhere (DMID 21-0004).^6^

### Immunogenicity

Binding immunoglobulin G (IgG) levels to full-length Spike (Spike) and to the receptor binding domain (RBD) of Spike evaluated using the validated Meso Scale Discovery (MSD) V-PLEX® SARS-CoV-2 Panel 2 IgG assay (MSD #K15383U)^7^ were bridged to international standards and reported as Binding Antibody Units (BAU/mL). SARS-CoV-2 neutralizing antibody (nAb) titers expressed as the serum inhibitory dilution required to achieve 50% neutralization (ID50) were evaluated by the live virus focus reduction neutralization titer (FRNT) assay with viruses representing SARS-CoV-2 Spike mutation D614G and Delta and Omicron (BA.1) variants as described previously.^8^ Detailed assay methods are in Supplementary Materials. Transplacental antibody transfer was evaluated by calculating the ratio of specific antibody levels in maternal and cord blood sera at the time of delivery.

### Statistical Analysis

Medians and interquartile ranges (IQRs) for binding IgG and ID50 nAb levels were summarized by study visit and vaccine type. Differences in antibody levels between groups at delivery were tested using regression analyses controlling for days since last vaccine dose, as well as sensitivity analyses that were restricted to participants with vaccination in the same time interval between last vaccination and delivery.

## Results

This report includes the first 240 pregnant participants who gave birth and their newborns: 100 Pfizer (102 infants) and 67 Moderna (68 infants) 2-dose vaccine recipients, and 73 booster dose participants (75 infants) (Table 1). Booster doses were mostly homologous with the primary series (80.8%). The median age of participants was 34 years (range, 22-51). Participants completed their primary 2-dose series at a median of 17.1 weeks of gestation, while booster vaccination was received at a median of 28.6 weeks of gestation. Post-vaccination sera were collected at a median of 18.7 (range: 1.6-33.3) weeks following completion of the 2-dose series and 6.0 (range: 1.1-19.9) weeks following the booster dose. The interval (median weeks) between last vaccine dose and delivery was shorter for booster dose recipients (10.4) than primary 2-dose recipients (21.7). Overall, 7.8% of primary 2-dose recipients and 16.4% of booster dose recipients had prior laboratory confirmed SARS-CoV-2 infection.

**Table 1.** Study Participant Characteristics.

| **Maternal Characteristic** | **Pfizer-BioNTech**<br>**n=100**<br>**Median (IQR)**<br>**% (n)** | **Moderna**<br>**n=67**<br>**Median (IQR)**<br>**% (n)** | **Booster**<br>**n=73**<br>**Median (IQR)**<br>**% (n)** |
| --- | --- | --- | --- |
| Age, years | 35 (31, 37) | 34 (31, 37) | 34 (31, 37) |
| Race |  |  |  |
| Asian | 12.0 (12) | 4.5 (3) | 9.6 (7) |
| Black/African American | 10.0 (10) | 16.4 (11) | 1.4 (1) |
| White | 75.0 (75) | 73.1 (49) | 83.6 (61) |
| Other | 3.0 (3) | 6.0 (4) | 5.5 (4) |
| Hispanic or Latino | 10.0 (10) | 14.9 (10) | 9.6 (7) |
| Vaccine Exposure up to Delivery |  |  |  |
| 2 doses Pfizer only | 98.0 (98) |  |  |
| 2 doses Pfizer+Pfizer boost | 2.0 (2) |  | 76.7 (56) |
| 2 doses Pfizer+Moderna boost | 0.0 (0) |  | 16.4 (12) |
| 2 doses Moderna only |  | 92.5 (62) |  |
| 2 doses Moderna+Moderna boost |  | 6.0 (4) | 4.1 (3) |
| 2 doses Moderna+Pfizer boost |  | 1.5 (1) | 2.7 (2) |
| Weeks Between Last Dose and Post-Vaccination Visit | 18.8 (14.6, 23.4) | 18.3 (12.9, 23.6) | 6.0 (3.3, 8.1) |
| Weeks Between Primary Series Completion and Delivery | 21.9 (18.0, 26.4) | 22.9 (18.3, 27.3) | 46.7 (40.4, 50.9) |
| Weeks Between Last Dose and Delivery | 21.6 (17.1, 25.7) | 21.9 (15.4, 24.6) | 10.4 (6.9, 14.1) |
| Gestational Age at First Dose During Pregnancy, weeks | 13.1 (9.3, 17.9) | 12.7 (7.6, 15.7) | -10.9 (-14.9, -4.6) |
| Gestational Age at Last Dose During Pregnancy, weeks | 16.9 (13.4, 21.3) | 17.1 (14.0, 23.7) | 28.6 (24.9, 31.7) |
| Gestational Age at Delivery, weeks | 39.3 (37.9, 40.1) | 39.1 (38.7, 39.9) | 39.1 (38.3, 39.6) |
| **Infant Characteristic** | **Pfizer-BioNTech**<br>**n=102**<br>**% (n)** | **Moderna**<br>**n=68**<br>**% (n)** | **Booster**<br>**n=75**<br>**% (n)** |
| Preterm birth (<37 weeks) | 6.9 (7) | 7.4 (5) | 14.7 (11) |
| Infant Birthweight g | 3370 (3040, 3640) | 3340 (3030, 3560) | 3280 (2860, 3540) |

### SARS-CoV-2 binding antibodies

Serum binding IgG to Spike and RBD were detected in all primary 2-dose and booster dose recipients at the post-vaccination and delivery visits, and in all cord blood samples (Figure 1). Significantly higher antibody levels were measured post-vaccination and at delivery in participants who received a booster vaccination during pregnancy compared to those who received only a primary 2-dose series. At delivery, the geometric mean titer (GMT) of IgG to Spike in booster vaccine recipients was 2,201 BAU/mL (n=73), 9.3-fold higher than in those receiving two doses of Pfizer (GMT 236 BAU/mL, n=100), and 4.6-fold higher than in those receiving two doses of Moderna (479 BAU/mL, n=67) vaccines (Figure 1).

**Figure 1.**
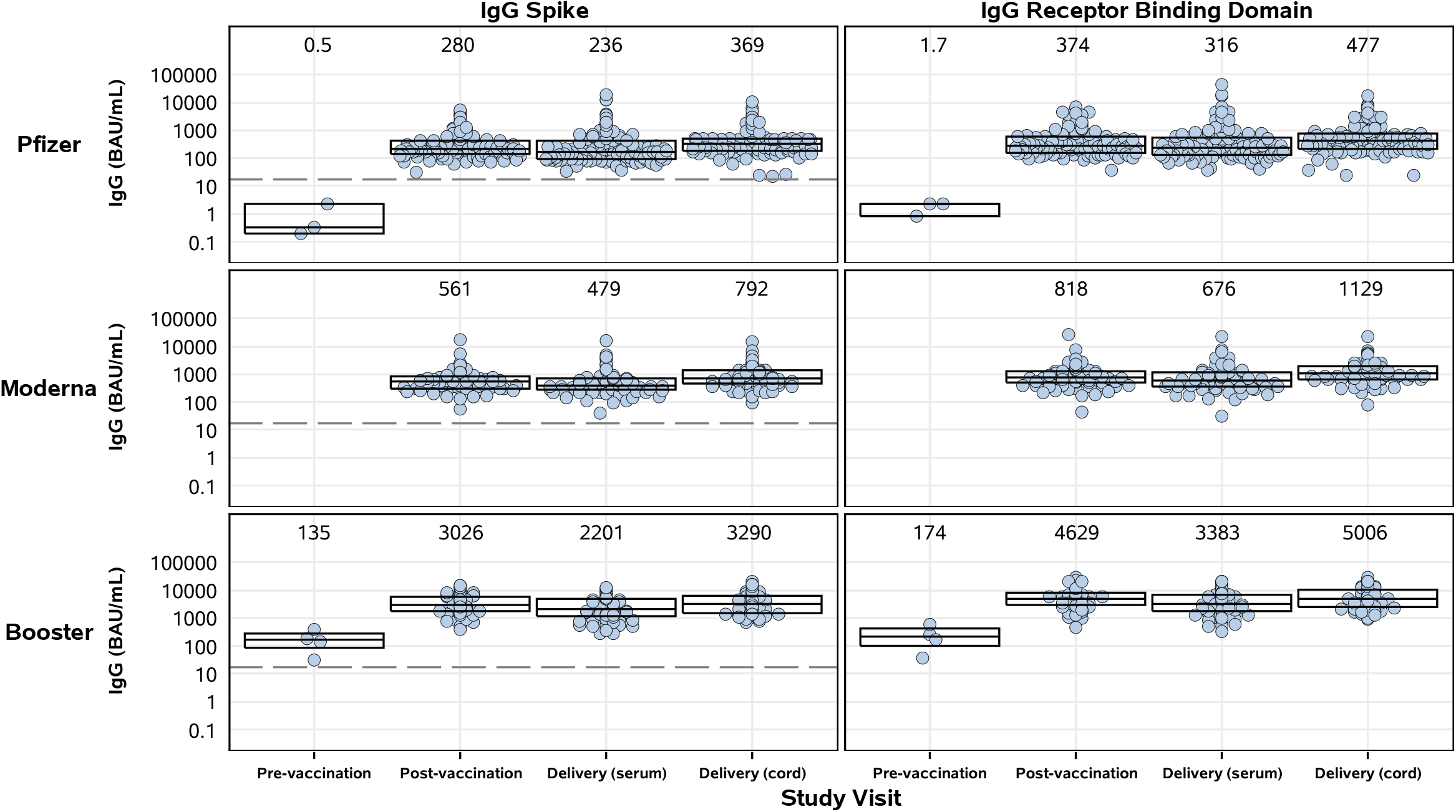
SARS-CoV-2 binding IgG in maternal and cord blood sera by study group and study visit. Pregnant participants received a 2-dose series of an mRNA vaccine (top – Pfizer, middle – Moderna) or a booster mRNA vaccine (bottom panel). Sera derived from maternal blood collected pre- and post-vaccination and at delivery, and cord blood, were evaluated for binding IgG to full-length Spike (left panels) and RBD (right panels). Titers were bridged to international standards and reported as Binding Antibody Units (BAU/mL). Box plots represent median (horizontal line within the box) and interquartile range; GMT is displayed at the top of each panel and the dashed line is the cutoff for positivity (17 BAU/mL).

Booster vaccination also elicited significantly higher levels of Spike IgG in cord blood, where the GMT was 3,290 BAU/mL, 8.9-fold and 4.2-fold higher than in cord blood from those vaccinated with two doses of Pfizer (GMT 369 BAU/mL) or Moderna (GMT 792 BAU/mL), respectively (Figure 1). Similar trends were observed for RBD IgG in cord blood and at the post-vaccination visit to both Spike and RBD IgG (Figure 1, Table S1).

Overall, the booster group and their infants had ∼0.6 log_10_ higher Spike and RBD IgG levels at delivery compared to the combined primary mRNA vaccine group (Pfizer and Moderna) after adjusting for days since last vaccination (p<0.0001) (Table S1). Sensitivity analyses showed similar results (data not shown).

### SARS-CoV-2 neutralizing antibodies

Significantly higher nAb titers to D614G were measured post-vaccination in pregnant participants who received a booster (GMT ID50 630.3) compared to those receiving a primary 2-dose series (GMT ID50 62.2 for Pfizer, 192.5 for Moderna) (Figure 2). High nAb titers persisted at delivery and were detectable in 100% of boosted participants (GMT ID50 446.4), compared to 68% (GMT ID50 49.9) and 96% (GMT ID50 179.5) of participants receiving 2 doses of Pfizer or Moderna, respectively (Figure 2, Tables S1, S2). While nAb titers to Omicron were present in only 9% (GMT ID50 12.7) of Pfizer and 22% (GMT ID50 14.7) of Moderna dosed participants at delivery, 73% (GMT ID50 60.2) of boosted participants had detectable nAb titers to Omicron (p<0.0001). Neutralizing activity against Delta was intermediate between D614G and Omicron.

**Figure 2.**
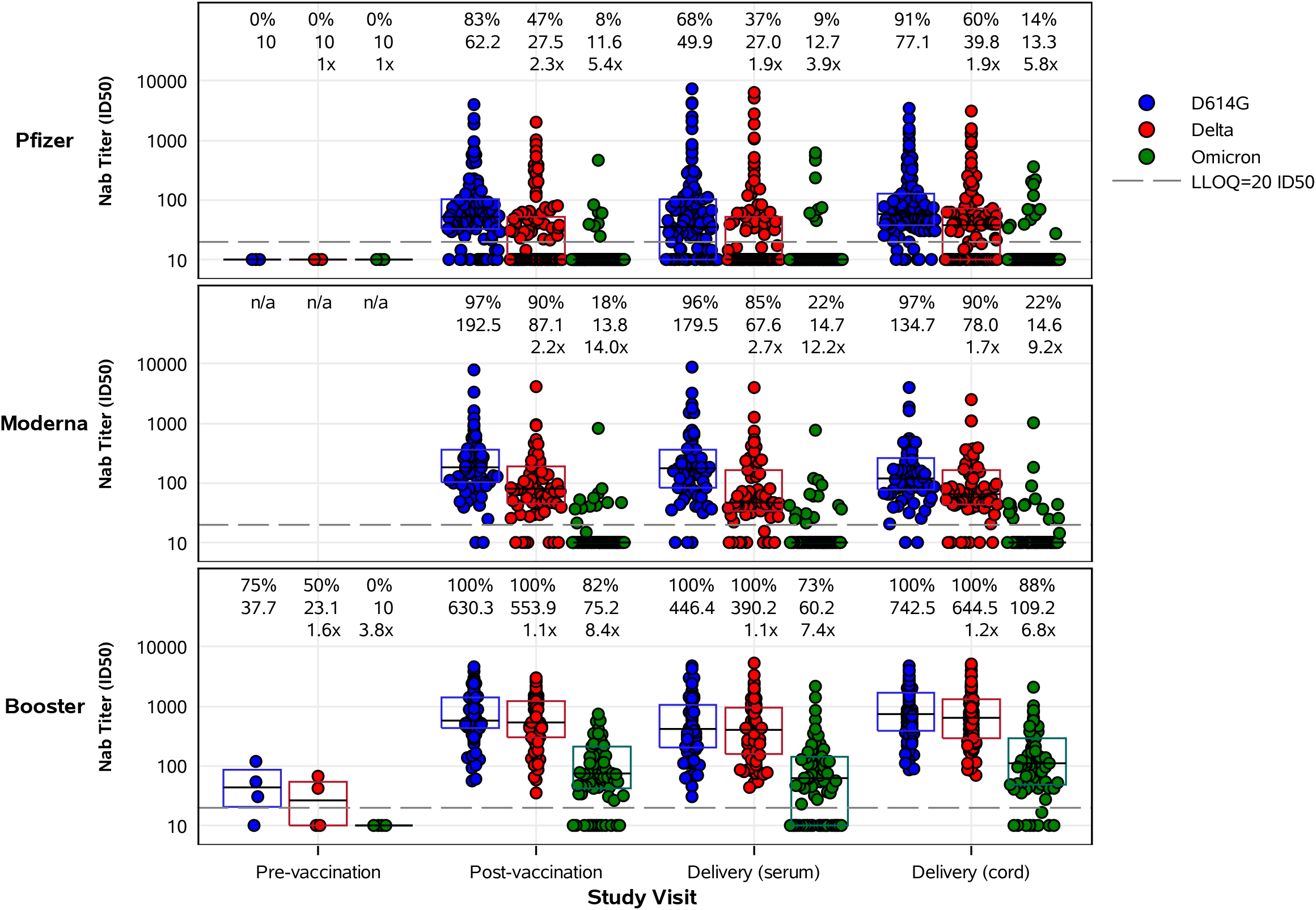
SARS-CoV-2 nAb activity of maternal and cord blood sera by study group and study visit. Pregnant participants received a 2-dose series of an mRNA vaccine (top – Pfizer, middle – Moderna) or a booster mRNA vaccine (bottom panel). Sera derived from maternal blood collected pre- and post-vaccination and at delivery, and cord blood, were evaluated for neutralization of D614G, Delta, and Omicron (BA.1) variants. Each point represents the GMT ID50 from two duplicates per specimen (within the same assay run). A value equivalent to half the lower limit of detection (LLOD = 20) was assigned to observations with no detectable response. A specimen was considered as having a positive response if at least one of the duplicates was above the LLOD. Box plots represent median (horizontal line within the box) and interquartile range. Response rate (% with responses > 20 ID50), GMT, and GMT fold reduction compared to D614G are displayed at the top of each panel.

Neutralizing antibody titers to D614G were also significantly higher in cord blood in the booster group (GMT ID50 742.5) compared to participants receiving 2 doses of Pfizer (GMT ID50 77.1) or Moderna (GMT ID50 134.7) (p<0.0001). Notably, nAb titers to Omicron were significantly higher in cord blood from the booster group (88% response rate, GMT ID50 109.2) compared to those receiving 2 doses of Pfizer (14% response rate, GMT ID50 13.3) or Moderna (22% response rate, GMT ID50 14.6) (p<0.0001). Sensitivity analyses showed similar results (data not shown).

### Transplacental antibody transfer

Efficient transplacental transfer was observed with both primary and booster vaccination during pregnancy, with median antibody transfer ratios between 1.55 and 1.77 for binding IgG and between 1.00 and 1.78 for nAb (Table S3).

## Discussion

In this large, multicenter observational cohort study, robust antibody responses to mRNA COVID-19 vaccines were detected in pregnant participants immunized across all gestational ages. The substantial increase in binding and neutralizing antibody titers measured in mothers and newborns at the time of delivery after a booster vaccination is a key finding which supports the administration of booster doses during pregnancy. This response was also observed in the nAb response to Omicron BA.1 where levels were significantly higher in booster recipients at delivery and in cord blood compared to those receiving a primary 2-dose series only. This finding is particularly relevant given the persistence of Omicron and its subvariants in this phase of the pandemic. However, the nAb levels to Omicron were significantly lower than to the vaccine-matched D614G variant, as expected and observed in non-pregnant populations.^9^ While the effect of prior maternal infection with SARS-CoV-2 on antibody responses, particularly against Omicron and in the booster vaccination group, cannot be precisely ascertained in this observational study, the proportion of previously infected mothers was small. This potential effect will be assessed in the full study cohort. Additionally, maternal binding IgG antibodies against both Spike and RBD SARS-CoV-2 proteins were efficiently transferred across the placenta and concentrated in the infant. This latter finding is particularly important given high hospitalization rates among infants <6 months old during the recent Omicron surge.^3^ Transplacental antibody transfer is a key component of newborn protection from SARS-CoV-2 infection and parallels demonstrated neonatal protection from respiratory pathogens such as influenza and pertussis.^10^

Given the observational design, the timing of vaccination during pregnancy was not pre-specified and the timing of sera collection post-vaccination was opportunistic. However, this investigation purposefully took a real-world scenario and inclusive enrollment approach and our analyses controlled for interval between vaccination and delivery to assess immune responses at the time of delivery. Thus, these data are generalizable and help delineate the potential impact of vaccination throughout pregnancy. Based on the effect of vaccination in non-pregnant populations, our study suggests likely heightened protection to mothers and newborns from booster doses during pregnancy.

Currently, pregnant women are not included among the risk groups targeted for a second booster dose of vaccine.^11^ While an absolute correlate of protection is still unknown, increases in antibody responses in adults have correlated with protection from symptomatic and severe COVID-19. Our findings support that COVID-19 mRNA vaccines should continue to be recommended during pregnancy with a clear emphasis on the sizable response to booster administration.

## Data Availability

Data collected for the study will be made available to others as a de-identified patient data set after finalization of clinical study report at the discretion of the IDCRC. Analyses of data, including data from staged analyses, will be available for presentation at scientific meetings and publication to inform the scientific community. If preliminary analyses are considered of public health importance or relevant to inform research, development, and implementation of SARS-CoV-2 vaccine in pregnancy, results may be shared with public health officials and partners to inform the global scientific community. The study will be conducted in accordance with the NIH Public Access Policy publication and data sharing policies and regulations. To request study data once complete, contact Flor M. Munoz

## List of Abbreviations

ACOG: American College of Obstetricians and Gynecologists
BAU/mL: Binding Antibody Units
CoVPN: COVID Prevention Network
DMID: Division of Microbiology and Infectious Diseases
DSMB: Data Safety monitoring Board
FRNT: Focus reduction neutralization titer
GMT: Geometric mean titer
IDCRC: Infectious Diseases Clinical Research Consortium
IgG: Immunoglobulin G
IQRs: Interquartile ranges
MSD: Meso Scale Discovery
nAb: Neutralizing antibodies
NIH: National Institutes of Health
RBD: Receptor binding domain
Spike: Full-length spike
U.S.: United States
VTEU: Vaccine Treatment and Evaluation Unit
WHO: World Health Organization

## Trial Status

This paper reflects Protocol Version 5.0, December 13, 2021; DMID 21-0004, ClinicalTrials.gov number <u>NCT05031468</u>. Recruitment began on July 6, 2021 and completed on January 31, 2022.

## Ethics Approval and Consent to Participate

Ethical approval of this protocol was received on May 28, 2021 by Vanderbilt University Medicine Center IRB, a single IRB as part of an NIH-funded consortium, IDCRC. Written informed consent was obtained from each participant.

## Acknowledgements

All the authors contributed to study design and have read and approved the final manuscript. We acknowledge Jeannie Murray, MS for contributions to the preparation and submission of this manuscript.

F.M.M. is an investigator of pediatric studies of COVID-19 vaccines for Pfizer and for a pediatric remdesivir study conducted by Gilead Sciences, Inc; serves as investigator on projects supported by an NIH contract for a Vaccine Treatment and Evaluation Unit (VTEU), serves as member of the Data Safety monitoring Board (DSMB) for clinical trials conducted by Pfizer, Moderna, Meissa Vaccines, Virometix, and the NIH; and is a member of the American Academy of Pediatrics Committee of Infectious Diseases (COID), the Immunization Expert Group of the American College of Obstetrics and Gynecology (ACOG), and Co-Chair of the COVAX Maternal Immunization Working Group.

K.M.N. is a member of the World Health Organization (WHO) Strategic Advisory Group of Experts on Immunization, serves as co-investigator on an NIH contract for a Vaccine Treatment and Evaluation Unit (VTEU), serves as Co-chair of the NIH COVID Prevention Network (CoVPN), and served as an investigator for Phase I/II Pfizer COVID-19 vaccine grant, with a grant to the institution, but no salary support. K.M.N. receives grants from Pfizer to conduct clinical trials of COVID vaccines through the Center for Vaccine Development and Global Health at University of Maryland, Baltimore. She receives grants from NIH to participate in overall organization of COVID-19 vaccine trials and for participation in vaccine trials.

M.J.M. conducts laboratory research and clinical trials with contract funding for vaccines or MABs vs SARS-CoV-2 with Lilly, Pfizer, and Sanofi and receives personal fees for Scientific Advisory Board service from Merck, Meissa Vaccines, Inc. and Pfizer.

M.S.S. served as an advisor for Moderna (ended December 2021) and is currently serving as an advisor for Ocugen, Inc.

B.A.R. currently holds a position on a DSMB for clinical trials at Gilead Sciences, Inc.

R.C.B. at Cincinnati Children’s Hospital receives research grant support for clinical trials from PATH, Astra Zeneca and Pfizer on which she serves as co-investigator.

B.B. owns shares in HDT Bio Corp.

J.S.G. receives research funds from NIH for Moderna KidCOVE study.

R.M.N. is a paid advisor to Gilead and an investigator on NIH-funded trials of Moderna, Pfizer and Janssen vaccines.

J.R-K is a medical speaker for Abbott Nutrition with the UIC team.

A.R.F. holds research grants from Pfizer, Janssen, Merck, Cyanvac, Biofire Diagnostics and serves on the DSMB for Novavax.

N.R. receives funds to conduct industry trials from Pfizer, Merck, and Sanofi-Pasteur and serves as a safety consultant for EMMES and ICON.

R.W.F. Jr., MD has received funds to conduct industry trials from Pfizer, Moderna and Astra Zeneca, serves on advisory boards for Merck, Sanofi-Pasteur, Johnson and Johnson and Seqirus and serves on an ICON-sponsored DSMB for a C difficile study.

All authors have completed relevant conflicts of interest in the Disclosure of Potential Conflicts of Interest section of the Authorship Form.

## Supplementary Appendix

This appendix is submitted by the authors to provide additional information about their work.

## Supplementary Appendix to Manuscript Entitled

**MASTHEAD**

The authors’ full names, academic degrees, email addresses by affiliations are as follows:

**Baylor College of Medicine (BCM), Houston, TX, Vaccine Research Center, Vaccine Treatment Evaluation Unit (VTEU)** (Protocol Chair)

Flor M. Munoz, MD, Departments of Pediatrics and Molecular Virology & Microbiology, Baylor College of Medicine, and Texas Children’s Hospital, Houston, TX 77030

**Emory University (The Hope Clinic of Emory University, Emory Children’s Center, Grady Memorial Hospital), Atlanta, GA, VTEU**

Martina L. Badell, MD, Department of Gynecology and Obstetrics, Division of Maternal Fetal Medicine, Emory University Hospital Midtown Perinatal Center, Atlanta, GA 30308

**University of Pittsburgh Medical Center (UPMC), Pittsburgh, PA, VTEU**

Katherine Bunge, MD, Department of Obstetrics, Gynecology and Reproductive Sciences, Magee-Women’s Hospital, Pittsburgh, PA 15213

**New York University (NYU) Langone Vaccine Center, New York, NY, VTEU**

*Manhattan Research Clinic:* Mark J. Mulligan, MD, NYU Langone Vaccine Center and Division of Infectious Diseases and Immunology, Department of Medicine, NYU Grossman School of Medicine, New York, NY 10016

*Brooklyn Research Clinic:* Lalitha Parameswaran, MD, NYU Langone Vaccine Center and Division of Infectious Diseases and Immunology, Department of Medicine, NYU Grossman School of Medicine, New York, NY 10016

**University of Rochester Medical Center, Rochester, NY, VTEU**

Courtney Olson-Chen, MD, Department of Obstetrics and Gynecology, University of Rochester, Rochester, NY 14642

**University of Illinois at Chicago, Chicago, IL, sub-site to Saint Louis University VTEU** Richard M. Novak, MD, Division of Infectious Diseases, University of Illinois, Chicago, IL 60612

**Cincinnati Children’s Hospital Medical Center (CCHMC), Cincinnati, OH, VTEU**

Rebecca C. Brady, MD, Cincinnati Children’s Hospital Medical Center, Division of Infectious Diseases, University of Cincinnati College of Medicine, Cincinnati, OH 45229

Emily DeFranco, DO, Department of Obstetrics and Gynecology, University of Cincinnati College of Medicine, Cincinnati, OH 45267

**Children’s Hospital of Philadelphia, Philadelphia, PA, sub-site to Vanderbilt University VTEU** Jeffrey S. Gerber, MD, PhD, Division of Infectious Diseases, Children’s Hospital of Philadelphia, University of Pennsylvania Perelman School of Medicine, Philadelphia, PA 19146

**Infectious Diseases Clinical Research Consortium (IDCRC) Laboratory Operations Unit (LOU)** Christine M. Posavad, PhD Vaccine and Infectious Disease Division, Fred Hutchinson Cancer Center; Department of Laboratory Medicine and Pathology, University of Washington, Seattle, WA 98109

**University of Maryland laboratory (binding Ab)**

Marcela Pasetti, PhD, Center for Vaccine Development and Global Health, University of Maryland School of Medicine, Baltimore, MD 21201

Mallory Shriver, MSc, Center for Vaccine Development and Global Health, University of Maryland School of Medicine, Baltimore, MD 21201

**Emory Vaccine Center laboratory (Live Virus Neutralization assay-FRNT)**

Mehul S Suthar, PhD Emory Vaccine Center, Yerkes National Primate Research Center; Department of Pediatrics; Department of Microbiology and Immunology, Emory School of Medicine, Emory University, Atlanta, GA 30322

Kathryn Moore, PhD, Emory Vaccine Center, Yerkes National Primate Research Center; Department of Pediatrics; Department of Microbiology and Immunology, Emory School of Medicine, Emory University, Atlanta, GA 30322

**Seattle Children’s Research Institute (Pseudovirus Neut)**

Rhea Coler, PhD, Seattle Children’s Research Institute, Center for Global Infectious Disease Research, Seattle, WA 98109

Bryan Berube, PhD, Seattle Children’s Research Institute, Center for Global Infectious Disease Research, Seattle, WA 98109

**IDCRC Leadership**

Kathleen M. Neuzil, MD, MPH, Center for Vaccine Development and Global Health, University of Maryland School of Medicine, Baltimore, MD 21201

**IDCRC Statistical and Data Science Unit (SDSU):**

Barbra A. Richardson, PhD, Departments of Biostatistics and Global Health, University of Washington, Vaccine and Infectious Disease Division, Fred Hutchinson Cancer Center, Seattle, WA 98109

**Statistical Center for HIV/AIDS Research and Prevention (SCHARP):**

Clifton Kelly, MS Statistical Center for HIV/AIDS Research and Prevention (SCHARP), Fred Hutchinson Cancer Center, Seattle, WA 98109

So Hee (Vicky) Kim, Statistical Center for HIV/AIDS Research and Prevention (SCHARP), Fred Hutchinson Cancer Center, Seattle, WA 98109

**Division of Microbiology and Infectious Diseases, National Institute of Allergy and Infectious Diseases, National Institutes of Health, Bethesda, MD**

Cristina Cardemil, MD, MPH

**National Institute of Allergy and Infectious Diseases, National Institutes of Health, Rockville, MD**

Jeanna M. Piper, MD

**FHI 360, Durham, NC**

Ashley Miller, MA, FHI 360, Durham, NC 27701

**University of Pittsburgh Medical Center (UPMC), Pittsburgh, PA, VTEU** (Protocol Chair)

Richard Beigi, MD, Department of Obstetrics, Gynecology and Reproductive Sciences, Magee-Women’s Hospital, Pittsburgh, PA 15213

**DMID 21-0004 Study Group**

**MOMI-Vax Study Group**

(listed in PubMed and ordered by lead institutions and enrollment)

The following study group members were all closely involved with the design, implementation, and oversight of the MOMI-Vax Study.

**Baylor College of Medicine (BCM), Houston, TX, Vaccine Training and Evaluation Unit (VTEU) (n=53)** (Protocol Chair)

Flor M. Munoz, MD, Departments of Pediatrics and Molecular Virology & Microbiology, Baylor College of Medicine, and Texas Children’s Hospital, Houston, TX 77030

Hana ElSahly, MD, Departments of Medicine and Molecular Virology and Microbiology, Baylor College of Medicine, Houston, TX 77030

Nanette Bond, PA-C, Department of Molecular Virology and Microbiology, Baylor College of Medicine, Houston, TX 77030

Patricia Santarcangelo, RN, Department of Molecular Virology and Microbiology, Baylor College of Medicine, Houston, TX 77030

Miguel Cantu, MS, Department of Molecular Virology and Microbiology, Baylor College of Medicine, Houston, TX 77030

**University of Pittsburgh Medical Center (UPMC), Pittsburgh, PA, VTEU (n=115)** (Protocol Chair) Richard Beigi, MD, Department of Obstetrics, Gynecology and Reproductive Sciences, Magee-Women’s Hospital, Pittsburgh, PA 15213

Katherine Bunge, MD, Department of Obstetrics, Gynecology and Reproductive Sciences, Magee-Women’s Hospital, Pittsburgh, PA 15213

**Emory University (Emory Clinic, Emory University Campus; The Hope Clinic of Emory Vaccine Center; The Hope Clinic of Emory Vaccine Center – Satellite Site; Emory Children’s Center; Emory Hospital Midtown; Emory Saint Joseph’s Hospital; Grady Memorial Hospital) (n=126 total across Emory locations)**

Martina L. Badell, MD, Department of Gynecology and Obstetrics, Emory University Hospital Midtown Perinatal Center, Atlanta, GA 30308

Nadine Rouphael, MD Hope Clinic of the Emory Vaccine Center, Division of Infectious Diseases, Department of Medicine, School of Medicine, Decatur, GA, USA

Anandi N. Sheth, MD, MSc Department of Medicine, Division of Infectious Diseases, Emory University School of Medicine, Grady Health System Atlanta, GA

Carolynn Dude MD, PhD, Department of Gynecology and Obstetrics, Division of Maternal Fetal Medicine, Grady Health System Atlanta, GA

**New York University Langone Vaccine Center, New York, NY, VTEU (Manhattan and Brooklyn Research Sites) (n=93 total across locations)**

Mark J. Mulligan, MD, NYU Langone Vaccine Center and Division of Infectious Diseases and Immunology, Department of Medicine, NYU Grossman School of Medicine, New York, NY 10016

Lalitha Parameswaran, MD, NYU Langone Vaccine Center and Division of Infectious Diseases and Immunology, Department of Medicine, NYU Grossman School of Medicine, New York, NY 10016

Ashley S. Roman, MD, MPH, Department of Obstetrics and Gynecology, NYU Langone Health, New York, NY 10016

Stephanie Sterling, MD, NYU Langone Vaccine Center and Division of Infectious Diseases and Immunology, Department of Medicine, NYU Grossman School of Medicine, New York, NY 10016

**University of Rochester Medical Center, Rochester, NY, VTEU (n=89)**

Courtney Olson-Chen, MD, Department of Obstetrics and Gynecology, University of Rochester, Rochester, NY 14642

Angela R. Branche, MD, Department of Medicine, Division of Infectious Diseases, University of Rochester, Rochester, NY 14642

Ann R. Falsey, MD, Department of Medicine, Division of Infectious Diseases, University of Rochester, Rochester, NY 14642

Erin Nowicki, Department of Medicine, Division of Infectious Diseases, University of Rochester, Rochester, NY 14642

**University of Illinois at Chicago, Chicago, IL, sub-site to Saint Louis University VTEU (n=54)** Richard M. Novak, MD, Division of Infectious Diseases, University of Illinois, Chicago, IL 60612

De-Ann Pillers, MD, Section of Pediatric Neonatology, University of Illinois, Chicago, IL 60612

Joann Romano-Keeler, MD, Section of Pediatric Neonatology, University of Illinois, Chicago, IL 60612

Alexis Braverman, MD, Department of Obstetrics and Gynecology, University of Illinois, Chicago, IL 60612

**Cincinnati Children’s Hospital Medical Center (CCHMC), Cincinnati, OH, VTEU (n=27)** Rebecca C. Brady, MD, Cincinnati Children’s Hospital Medical Center, Division of Infectious Diseases, University of Cincinnati College of Medicine, Cincinnati, OH 45229-3039

Emily DeFranco, DO, Department of Obstetrics and Gynecology, University of Cincinnati College of Medicine, Cincinnati, OH 45267

Robert W. Frenck, Jr., MD, Cincinnati Children’s Hospital Medical Center, Division of Infectious Diseases, University of Cincinnati College of Medicine, Cincinnati, OH 45229-3039

Donna Cunha, RN, Cincinnati Children’s Hospital Medical Center,

Division of Infectious Diseases, University of Cincinnati College of Medicine, Cincinnati, OH 45229-3039

Madison Minette Department of Obstetrics and Gynecology, University of Cincinnati College of Medicine, Cincinnati, OH 45267

**Children’s Hospital of Philadelphia, Philadelphia, PA, sub-site to Vanderbilt University VTEU (n=18)**

Jeffrey S. Gerber, MD, PhD, Division of Infectious Diseases, Children’s Hospital of Philadelphia, University of Pennsylvania Perelman School of Medicine, Philadelphia, PA 19146

Karen Puopolo, MD, PhD, Division of Neonatology, Children’s Hospital of Philadelphia, University of Pennsylvania Perelman School of Medicine, Philadelphia, PA 19146

Dustin Flannery, DO, MSCE, Division of Neonatology, Children’s Hospital of Philadelphia, University of Pennsylvania Perelman School of Medicine, Philadelphia, PA 19146

Krisha Patel, MS, Children’s Hospital of Philadelphia, Philadelphia, PA 19146

**Infectious Diseases Clinical Research Consortium (IDCRC) Laboratory Operations Unit (LOU)** Christine M. Posavad, PhD Vaccine and Infectious Disease Division, Fred Hutchinson Cancer Center; Department of Laboratory Medicine and Pathology, University of Washington, Seattle, WA

Weston Lawler, Vaccine and Infectious Disease Division, Fred Hutchinson Cancer Center, Seattle, WA

**University of Maryland laboratory (binding Ab)**

Marcela Pasetti, PhD, Center for Vaccine Development and Global Health, University of Maryland School of Medicine, Baltimore, MD

Mallory Shriver, MSc, Center for Vaccine Development and Global Health, University of Maryland School of Medicine, Baltimore, MD

Cheilon Bolanos, BA, Center for Vaccine Development and Global Health, University of Maryland School of Medicine, Baltimore, MD

Jennifer Oshinsky, BS, Center for Vaccine Development and Global Health, University of Maryland School of Medicine, Baltimore, MD

**Emory Vaccine Center laboratory (Live Virus Neutralization assay-FRNT)**

Mehul S Suthar, PhD Emory Vaccine Center, Yerkes National Primate Research Center; Department of Pediatrics; Department of Microbiology and Immunology, Emory School of Medicine, Emory University, Atlanta, GA

Kathryn Moore, PhD, Emory Vaccine Center, Yerkes National Primate Research Center; Department of Pediatrics; Department of Microbiology and Immunology, Emory School of Medicine, Emory University, Atlanta, GA

Kelly E. Manning, MS, MPH, Emory Vaccine Center, Yerkes National Primate Research Center; Department of Pediatrics; Department of Microbiology and Immunology, Emory School of Medicine, Emory University, Atlanta, GA

Alberto Moreno, MD, Emory Vaccine Center, Yerkes National Primate Research Center; Department of Pediatrics; Department of Microbiology and Immunology, Emory School of Medicine, Emory University, Atlanta, GA

**Seattle Children’s Research Institute (Pseudovirus Neut)**

Rhea Coler, PhD, Seattle Children’s Research Institute, Seattle, WA 98109

Bryan Berube, PhD, Seattle Children’s Research Institute, Seattle, WA 98109

**IDCRC Leadership**

Kathleen M. Neuzil, MD, MPH, Center for Vaccine Development and Global Health, University of Maryland School of Medicine, Baltimore, MD 21201

**IDCRC Statistical and Data Science Unit (SDSU):**

Barbra A. Richardson, PhD, Departments of Biostatistics and Global Health, University of Washington, Vaccine and Infectious Disease Division, Fred Hutchinson Cancer Center, Seattle, WA

**Statistical Center for HIV/AIDS Research and Prevention (SCHARP):**

Clifton Kelly, MS, Statistical Center for HIV/AIDS Research and Prevention (SCHARP), Fred Hutchinson Cancer Center, Seattle, WA

So Hee (Vicky) Kim, Statistical Center for HIV/AIDS Research and Prevention (SCHARP), Fred Hutchinson Cancer Center, Seattle, WA

Lauren Young, MPH, Statistical Center for HIV/AIDS Research and Prevention (SCHARP), Fred Hutchinson Cancer Center, Seattle, WA

Wen-Min (Wendy) Hou, MPH, BSN, CCRP, Statistical Center for HIV/AIDS Research and Prevention (SCHARP), Fred Hutchinson Cancer Center, Seattle, WA

**Division of Microbiology and Infectious Diseases, National Institute of Allergy and Infectious Diseases, National Institutes of Health, Bethesda, MD**

Cristina Cardemil, MD, MPH

**National Institute of Allergy and Infectious Diseases, National Institutes of Health, Rockville, MD**

Jeanna M. Piper, MD

**FHI 360, Durham, NC**

Ashley Miller, MA, FHI 360, Durham, NC

**MOMI-Vax Study Team Members**

The MOMI-Vax study was a collaborative effort at many sites within the Infectious Diseases Clinical Research Consortium. Below is a list of sites and staff that worked tirelessly to implement and conduct the MOMI-Vax study:

**Baylor College of Medicine (BCM), Houston TX, VTEU**

Flor M. Munoz, MD, Nanette Bond PA-C, Patricia Santarcangelo RN, Miguel Cantu, MS, Erin Nicholson, MD, Kjersti Aagaard, MD, PhD, Tony Piedra, MD, Hana ElSahly, Yvette Rugeley, Lisreina Toro, Connie Rangel, Tina Sierra, Vanessa Martinez, Kathy Bosworth, Yolanda Rayford, Marinna Matta, Keely Wilson, Tykel Eddy, Jesus Banay, Kayla Burell, Jeremy Castro, Chianti Wade-Bowers, Velicia Delane, Rachel Saenz, RN, Hoyt L Hoffman, RN

**University of Pittsburgh Medical Center (UPMC), Pittsburgh, PA, VTEU**

Richard Beigi, MD, Katherine Bunge, MD, Jamie Haggerty, Ingrid Macio, Danielle Litzinger, Tationna Smalley, Tracy Campbell, Anne-Marie Rick

**Emory University (The Hope Clinic of Emory Vaccine Center – Satellite Sites: Emory Children’s Center; Emory Hospital Midtown; Grady Memorial Hospital, Emory Clinic), Atlanta, GA, VTEU** Martina L. Badell, MD, Nadine Rouphael, MD, Anandi Sheth MD, Colleen Kelley MD, Valeria Cantos MD, Michael Chung MD, Paulina Rebolledo MD, Caitlin Anne Moran, MD, Carolynn Marie Dude, MD PhD, Brittany Spiegel, NP-C, Christina Blanca Bacher, PA-C, Pamela Weizel, C-FNP, Erica Baker, RN, Riaun Floyd, Juliet Brown, Chris Foster, Les’Shon Irby, Christin Root, Arelis Villot Santiago, Cassie Grimsley-Ackerley, MD, Veronica Smith, NP, Jessica Traenkner, PA, Sarah Bechnak, RN, Deborah Laryea, RN, Hollie Macenczak, RN, Amer Bechnak, MD, Lana Khalil, MD, Eduardo Monarrez, Terra Jean Winter, Evan Gutter, Lisa Harewood, Dilshad Rafi Ahmed, Alahna Watson, Brittany Robinson, Brandi Johnson, Yongxian Xu, MD, Natalie Gray, Christopher Huerta, Bernadine Panganiban, Jacob Usher, Dongli Wang, Michele McCullough, Vanessa Soliman, RN, Stephanie Ramer, Cecilia Zhang, Ariel Kay, Ghina Alaeddine, MD, Laurel Bristow, Amal Naji, Mai Kio, Stacia Crochet MD, Kareem Bechnak, RN, Tolulope Ojo-Akosile, Mary Atha, NP, Kristen Unterberger, PA, Evan Anderson, MD, Ash Grimes, Mindy Fletcher

**New York University Langone Vaccine Center, New York, NY, VTEU**

*Manhattan Research Site:*

Mark Mulligan, MD, Ashley S. Roman, MD, MPH, Ashanay Allen, Tamia Davis, NP, Celia Engelson, NP, Stephanie Rettig, Pamela Suman, Madalyn Saporito, Marie Samanovic-Golden, PhD, Heekoung Youn, RN, Thomas Jenkins, RN, Abdonnie Holder, Samantha Yip, RN, Meron Tasissa, Jimmy Wilson, Shelby Goins, Jacqueline Callahan, RN, Ramin Herati, MD, Alexander McMeeking, MD, Purvi Parikh, MD, Rebecca Pellett Madan, MD, Bo Shopsin, MD, Vijaya Soma, MD, Vanessa Raabe, MD, Lisa Zhao, Mala Begum, Manhoor Ali, Rachel Briks, RN, Juanita Erb, RN, Tiairah McNeill, Nicholas Renton, Angelica Kottkamp, MD, Edward Nirenberg, Natella Aronova, NP

*Brooklyn Research Site:*

Lalitha Parameswaran, MD, Stephanie Sterling, MD, Miguel Rodriguez, Emilie Geesey, Daniel Sartori, MD, Kate Baicy, MD, Grace Lioue, NP, Thomas Jenkins, RN, Andrew Korman, Akeem Moore, Heycha Rosado-Lebron, Diandra Williams, Jasmine Briscoe, Kelly Minus, NP, Nora Henderson, Paula Tamashiro Tairaku, MD

**University of Rochester Medical Center, Rochester, NY, VTEU**

Courtney Olson-Chen, MD, Angela Branche, MD, Ann Falsey, MD, Mary Caserta, MD, Cynthia Rand, MD Doreen Francis, RN, Erin Nowicki, HPC, Ian Shannon, RN, Sophia Wiltse, HPC, Sharon Moorehead, HPC, Michael Peasley, Kari Steinmetz HSRC, Katherine Elena, RN Elizabeth Barker, RN Tanya Smith, HSRC, Samuel

Diehl, Amy Jasek, HPC, Sarah Cox, HPC, Tumininu Faniyan, William Hamilton, Sarah Caveglia, HPC, Haley Burger, Christopher Lane, Arthur Zemanek, RN, Spencer Obrecht, RN, Patrick Kingsley, HPC

**UIC**

Richard M. Novak, MD, Andrea Wendrow, RPh, Stephanie Martin, RN, Alexis Braverman, MD, Caitlin Buhimschi, MD, Julie Hartwig, RN, De-Ann Pillers,MD, Zaynab Kadhem, Desmona Strahan, Karen Hayani,MD, Joann Romano-Keeler,MD, Habiba Sultana, Nanu Kunwar, Khandaker Anwar, Mahmood Ghassemi, Md-Ruhul Amin, Braulio Carrasco, Tasmin Sultana, Henna Park, MD, Vanessa Orlando Castellano, Wilhelmina Brown

**Cincinnati Children’s Hospital Medical Center (CCHMC), Cincinnati, OH, VTEU**

Rebecca Brady, MD, Kristen Buschle, Robert W. Frenck, Jr., MD, Jamie Kidd, Jesse LePage, Sarah McCartney, Nicole Meyer, Laura Pace, Grant Paulsen, Stacy Ranz, Paul Spearman, Jennifer Whitaker, Eleanor Widdice, Felicia Scaggs-Huang, Haley Muth, Susan Parker, RN, Michelle Dickey, APRN, Donna Cunha, Hannah Ingraham, Vivian Mulholland, Tena Pham, Julie Kulhanek, Yislain Villalona, Beth Higgins, Lloyd Zoellner, Theresa Baker, Eric Leon, Mary Pat McKee, Marian Crossman, RN, Monica McNeal, Teresa Mastin-Diebold, Kendall Comeaux, Courtney Budke, Natalie Brinkmann, Tammy Lewis-McCauley, Behroze Dalal

***University of Cincinnati Medical Center, Cincinnati, OH, sub-site of CCHMC VTEU***

Emily DeFranco, DO, Madison Minette

**CHOP**

Jeffrey S. Gerber, MD, PhD, Karen Puopolo, MD, PhD, Dustin Flannery, DO, MSCE, Rasheeda Lawler, MPH, BSN, RN, Toni Mancini, RN, CCRC, Tevin Carrington, Krisha Patel, MS, Emily Woodford, Madeline Pfeifer, Richard Tustin, Annemarie Butler, Shawn O’Connor, Mahaa Ahmed, Maya Wright, Stephanie Tack, Minha Sarwar, Molly Tancini, Audrey Kamrin

**IDCRC Laboratory Operations Unit (LOU), Vaccine and Infectious Disease Division, Fred Hutchinson Cancer Center**

Christine M. Posavad, PhD, Weston Lawler, Michael Stirewalt, Megan Meagher

**Center for Vaccine Development and Global Health, University of Maryland School of Medicine, Baltimore, MD (binding Ab)**

Marcela Pasetti, PhD, Mallory Shriver, MSc, Cheilon Bolanos, BA, Jennifer Oshinsky, BS

**Emory Vaccine Center laboratory (Live Virus Neutralization assay-FRNT)**

Mehul S Suthar, PhD, Kathryn Moore, PhD, Kelly E. Manning, MS, MPH, Alberto Moreno, MD, Madison Ellis

**Seattle Children’s Research Institute (Pseudovirus Neut)**

Rhea Coler, PhD, Bryan Berube, PhD

**IDCRC Statistical and Data Science Unit (SDSU), Vaccine and Infectious Disease Division, Fred Hutchinson Cancer Center, Seattle, WA:**

Barbra A. Richardson, PhD, Elizabeth R. Brown, ScD, Jen Balkus, PhD

**Statistical Center for HIV/AIDS Research and Prevention (SCHARP), Fred Hutchinson Cancer Center, Seattle, WA:**

Clifton Kelly, MS, So Hee (Vicky) Kim, Lauren F. Young, MPH, Wen-Min (Wendy) Hou, MPH, BSN, CCRP, Karan A. Shah, MS, Udhab Adhikari, PhD, Craig N. Chin, MS, Anisa C. Gravelle, MS, Hannah

Hale, Tanya Harrell, Yun Ji (Nina) Lee, CCRP, Melissa J. Simon, Chloe Waters, MPH, MSW, Josh Larkin, DASM, MSc

**IDCRC Leadership and Administrative Team**

Kathleen M. Neuzil, MD, David S. Stephens, MD, Monica M. Farley MD, Jeanne Marrazzo, MD, Robert

L. Atmar MD, Jeffery Lennox, MD, Sidnee Paschal Young, Barbara E. Walsh

**Division of Microbiology and Infectious Diseases, National Institute of Allergy and Infectious Diseases, National Institutes of Health, Bethesda, MD**

Cristina Cardemil, MD, MPH, Olivia Sparer, Marina Lee, PhD, Seema Nayak, MD

**National Institute of Allergy and Infectious Diseases, National Institutes of Health, Rockville, MD**

Jeanna Piper, MD

**Biomedical Advanced Research & Development Authority, U.S. Department of Health and Human Services (DHHS), Washington, DC**

Mirjana Nesin, MD, FEEP

**FHI 360, Durham, NC**

Ashley Miller, MA, Katlyn Hurst, Anna R. Borough, MSc, Joy Miedema, MPH, Linda McNeil, MA, Mary Briggs, Beth Horn

## Supplemental Methods

### Supplemental Methods 1: SARS-CoV-2 Binding Antibody Assay Methods

The humoral immune responses to SARS-CoV-2 vaccination and durability of vaccine-induced antibodies (Ab) in pregnant women and their infants were assessed with a quantitative, high throughput multiplexed binding antibody assay. Serum IgG against SARS-CoV-2 Spike protein, and S1 receptor binding domain (RBD) were measured using the Meso Scale Discovery (MSD) V-PLEX® SARS-CoV-2 Panel 2 IgG assay (MSD #K15383U) according to manufacturer’s instructions. The MSD V-PLEX assays have been validated^1^ by the manufacturer and have been employed in numerous studies to evaluate antibody responses to SARS-CoV-2 infection and/or vaccination.^1-7^

MSD multiplex assays function similarly to a traditional ELISA; however, multiple antigens are bound to distinct spots in a single well of a 96-well plate, thereby allowing for the simultaneous measurement of specific Ab against multiple antigens. Assays were performed according to manufacturer’s instructions. In brief, SARS-CoV-2 Plate 2 (MSD #N05380A-1) assay plates, MSD Blocker A solution (MSD #R93AA-2), and Diluent 100 (MSD #R50AA-3) were removed from 4°C storage and allowed to acclimate to room temperature (RT). Reference Standard 1 (MSD #C00ADK-2), Serology Controls (MSD #C00ADK-2), and serum samples were removed from -80°C storage, thawed on ice, and allowed to come to RT. Assay plates were blocked by adding 150 μL/well of Blocker A for 30-120 mins on microplate shaker (700 rpm) in RT incubator (22°C). After blocking, plates were washed three times with 150 μl/well 1X phosphate buffered saline and 0.05% Tween-20 (PBS-T). Serum samples were initially diluted to 1:5,000 in Diluent 100 and added to plates in duplicate (50 μL/well). Serology Controls 1.1, 1.2, and 1.3 were added undiluted to each plate in duplicate (50 μL/well). Seven-point Calibrators and a blank (Diluent 100 only) were prepared from dilution of Reference Standard 1, diluted 1:10 in Diluent 100 and 4-fold serially diluted. Calibrators were included in duplicate (50 μL/well) on every plate. Plates were incubated for 2 h with 700 rpm shaking in RT incubator and subsequently washed with PBS-T as described above. Ab were detected with a conjugated SULFO-TAG™ Anti-Human IgG Antibody (MSD #D21ADF-3) diluted (1X) in Diluent 100. Detection Ab was added at 50 μL/well to all wells. Plates were again incubated for 1 h with 700 rpm shaking in RT incubator and were then washed with PBS-T. MSD GOLD Read Buffer B (MSD #R60AM-3) was added to plates (150 μL/well), and plates were immediately read on the MSD QuickPlex SQ 120 reader. The instrument discharged an electrical current through the plate and measured the electrochemiluminescence (ECL) signal emitted by the bound detection Ab on each distinct antigen.

The raw data (ECL signals) were analyzed using MSD Discovery Workbench Version 4.0 software, which automatically subtracted the mean ECL signal of the blanks from sample data to generate adjusted ECL signals. The Workbench software generated a standard curve for each antigen by fitting the ECL signals from the 7-point calibrations curve (i.e., serial dilution of Reference Standard 1) to a four-parameter logistic (4PL) regression model. Ab concentrations in arbitrary units (AU)/mL of the Serology Controls and diluted sera were determined by the Workbench software. ECL signals were backfitted to the standard curve, and final Ab concentrations in undiluted samples were dilution corrected (calculated Ab concentration x sample dilution factor). The mean Ab result (AU/mL) of duplicates was reported.

Prior to testing IDCRC 21-0004 samples, quality control (QC) parameters and acceptance criteria were established, and an in-house standard operating procedure (SOP) was prepared by UMB. Goodness of fit (R2) of the 4PL regression was assessed in Workbench: for an assay to be accepted R2 was ≥ 0.98. Additionally, the %Recovery of each point in each standard curve was examined. Acceptable %Recovery was defined as 80-120%. The acceptable ranges of the Serology Controls were defined as ± 20% of the known Ab concentrations (Ab concentrations in AU/mL provided by MSD). The maximum variation of calculated Ab concentrations allowed between duplicates in standards or controls was a coefficient of variation (%CV) ≤ 20%. If the standards or controls did not satisfy these criteria, then an assay or an entire plate was repeated. The mean ECL signal of the blanks was required to be < 200 otherwise the assay was repeated.

For serum samples, if the %CV between duplicates was >25%, the test was repeated. If the adjusted ECL signal of a sample was outside of the assay range (automatically determined by Workbench), then the sample was retested for that assay at an appropriate dilution. If the adjusted ECL signal was above the upper limit of quantification (ULOQ), the sample was repeated at a higher (less concentrated) dilution. Conversely, if the signal was below the lower limit of quantification (LLOQ) for an assay, the sample was repeated using a lower (more concentrated) dilution. ULOQ and LLOQ for each assay were tabulated by multiplying the upper and lower limits of detection (LOD) (determined by the manufacturer) by the lowest (1:100) and highest (1:100,000) sample dilutions allowed, respectively. If all plate and sample QC criteria were satisfied, the mean calculated Ab concentration (AU/mL) was reported for a sample. Additionally, the results were reported in WHO international units. Reference Standard 1 was calibrated against the WHO International Standard (NIBSC code: 20/136) by the manufacturer and a conversion factor for each antigen was provided to allow for conversion from MSD AU/mL to WHO/NIBSC binding antibody units (BAU/mL).

Additionally, a positivity cut-off for SARS-CoV-2 spike IgG in serum was established. Spike IgG levels were measured in historic sera (i.e., collected prior to the emergence of SARS-CoV-2, n=49) and post-COVID vaccination serum samples (n=183) with MSD V-PLEX Panel 2 as described above. Using a receiver operating characteristic (ROC) curve, S IgG (BAU/mL) and associated variables were analyzed. A Spike IgG cut-off value of 17 BAU/mL (resulting in 100% specificity and sensitivity to detect positive samples) was established.^8^ A similar positivity cut-off level has been reported in the literature.^1,9,10^

### Supplemental Methods 2: Live Virus Focus Reduction Neutralization (FRNT) Assay

#### Cell Lines

VeroE6-TMPRSS2 cells used in the FRNT assay were generated and cultured as previously described.^11^Viruses. For the FRNT assay, the EHC-083E (SARS-CoV-2/human/USA/GA-EHC-083E/2020) variant was isolated and propagated as previously described.^12^ hCoV-19/USA/PHC658/2021 (herein referred to as the B.1.617.2 variant or Delta) was derived from a nasal swab collected in May 2021.^11^ hCoV19/EHC_C19_2811C (referred to as the B.1.1.529 variant or Omicron) was derived from a mid-turbinate nasal swab collected in December 2021. This SARS-CoV-2 genome is available under GISAID accession number EPI_ISL_7171744. All viruses used in the FRNT assay were deep sequenced and confirmed as previously described.^11^

| Live virus variant | Spike mutations |
| --- | --- |
| EHC-083E | D614G |
| Delta B.1.617.2 | T19R, K77T, G142D, Del (E156-R158); Ins G, L452R, T478K, D614G, P681R, D950N |
| Omicron (BA.1) | A67V, Δ69-70, T95I, G142D, Δ143-145, Δ212, N211I, +214EPE, G339D, S371L, S373P, S375F, K417N, N440K, G446S, S477N, T478K, E484A, Q493R, G496S, Q498R, N501Y, Y505H, T547K, D614G, H655Y, N679K, P681H, N764K, D796Y, N856K, Q954H, N969K, L981F |

FRNT assays were performed as previously described.^6,11,13^ Briefly, samples were diluted at 3-fold in 8 serial dilutions using DMEM in duplicates with an initial dilution of 1:10 in a total volume of 60 μl. Serially diluted samples were incubated with an equal volume of D614G, B.1.617.2 or B.1.1.529 (100-200 foci per well based on the target cell) at 37º C for 45 minutes in a round-bottomed 96-well culture plate. The antibody-virus mixture was then added to VeroE6-TMPRSS2 cells and incubated at 37º C for 1 hour. Post-incubation, the antibody-virus mixture was removed and 100 μl of pre-warmed 0.85% methylcellulose (Sigma-Aldrich, #M0512-250G) overlay was added to each well. Plates were incubated at 37º C for either 18 hours (D614G, B.617.2) or 40 hours (B.1.1.529) and the methylcellulose overlay was removed and washed six times with PBS. Cells were fixed with 2% paraformaldehyde in PBS for 30 minutes. Following fixation, plates were washed twice with PBS and permeabilization buffer (0.1% BSA and 0.1% Saponin in PBS) was added to permeabilized cells for at least 20 minutes. Cells were incubated with an anti-SARS-CoV spike primary antibody directly conjugated to Alexaflour-647 (CR3022-AF647) for 4 hours at room temperature or overnight at 4ºC. Cells were washed three times in PBS and foci were visualized on an ELISPOT reader. Antibody neutralization was quantified by counting the number of foci for each sample using the Viridot program.^14^ The neutralization titers were calculated as follows: 1 - (ratio of the mean number of foci in the presence of sera and foci at the highest dilution of respective sera sample). Each specimen was tested in duplicate. The FRNT-50 titers were interpolated using a 4-parameter nonlinear regression in GraphPad Prism 9.2.0. Samples that do not neutralize at the limit of detection at 50% are plotted at 10 and was used for geometric mean and fold-change calculations.

#### Unit of analysis

Neutralization titers reported as below the lower limit of detection (LLOD) were assigned a value equivalent to half the LLOD. For FRNT, the unit of analysis was obtained as the Geometric Mean of the duplicate titers. Positive response (at the sample level) was defined as titers above the LLOQ or at least one of the duplicate titers above the LLOQ.

## Supplemental Tables

**Table S1.** Differences in Antibody Levels Between Primary 2-Dose and Booster Groups at Delivery.

| Lab Assay | Sample | Primary GMT<br>(95% CI) | Booster<br>GMT<br>(95% CI) | Mean Log<sub>10</sub><br>Difference*<br>(95% CI) | p-value |
| --- | --- | --- | --- | --- | --- |
| Spike IgG BAU/mL | Maternal<br>Delivery | 313.8<br>(260.7, 377.8) | 2200.6<br>(1764.9, 2743.8) | 0.58<br>(0.44, 0.71) | <0.0001 |
|  | Cord<br>Blood | 500.5<br>(424.1, 590.7) | 3290.4<br>(2713.6, 3989.9) | 0.59<br>(0.46, 0.72) | <0.0001 |
| RBD IgG BAU/mL | Maternal<br>Delivery | 428.6<br>(351.9, 522.1) | 3382.9<br>(2703.7, 4232.86) | 0.62<br>(0.47, 0.77) | <0.0001 |
|  | Cord<br>Blood | 673.1<br>(564.5, 802.6) | 5005.7<br>(4126.5, 6072.1) | 0.64<br>(0.50, 0.78) | <0.0001 |
| D614G nAb ID50 | Maternal<br>Delivery | 83.4<br>(65.4, 106.4) | 446.4<br>(342.1, 582.5) | 0.48<br>(0.29, 0.67) | <0.0001 |
|  | Cord<br>Blood | 96.4<br>(80.2, 115.9) | 742.5<br>(588.5, 936.8) | 0.69<br>(0.53, 0.84) | <0.0001 |
| Delta nAb ID50 | Maternal<br>Delivery | 39.0<br>(30.9, 49.2) | 390.2<br>(299.2, 508.9) | 0.78<br>(0.59, 0.97) | <0.0001 |
|  | Cord<br>Blood | 52.1<br>(42.3, 64.2) | 644.5<br>(507.5, 818.4) | 0.88<br>(0.71, 1.05) | <0.0001 |
| Omicron nAb ID50 | Maternal<br>Delivery | 13.5<br>(11.9, 15.3) | 60.2<br>(43.6, 83.1) | 0.55<br>(0.40, 0.69) | <0.0001 |
|  | Cord<br>Blood | 13.8<br>(12.2, 15.6) | 109.2<br>(81.6, 146.1) | 0.81<br>(0.68, 0.94) | <0.0001 |
| Pseudotyped WT nAb<br>ID50 | Maternal<br>Delivery | 93.4<br>(77.3, 112.9) | 512.5<br>(399.1, 658.3) |  |  |
|  | Cord<br>Blood | 220.8<br>(183.5, 265.7) | 941.4<br>(735.7, 1204.7) |  |  |

\*From regression analysis adjusted for days since last vaccination prior to delivery.

**Table S2.** Differences in Response Rate for nAb Between Primary 2-Dose and Booster Groups at Delivery.

| Lab Assay | Sample | Primary Response Rate* | Booster Response Rate* | Prevalence Ratio** (95% CI) | p-value |
| --- | --- | --- | --- | --- | --- |
| D614G nAb | Maternal Delivery | 79.0% (132/167) | 100% (73/73) | n/a*** |  |
|  | Cord Blood | 93.5% (159/170) | 100% (75/75) | n/a*** |  |
| Delta nAb | Maternal Delivery | 56.3% (94/167) | 100% (73/73) | n/a*** |  |
|  | Cord Blood | 71.8% (122/170) | 100% (75/75) | n/a*** |  |
| Omicron nAb | Maternal Delivery | 14.4% (24/167) | 72.6% (53/73) | 2.25 (1.66, 3.06) | <0.0001 |
|  | Cord Blood | 17.1% (29/170) | 88.0% (66/75) | 2.09 (1.66, 2.65) | <0.0001 |
| Pseudotyped WT nAb |  |  |  |  |  |

\*Response rate defined as number of samples above the lower limit of detection divided by number of samples tested.
\*\*From log-binomial regression analysis adjusted for days since last vaccination prior to delivery.
\*\*\*Model did not converge.

**Table S3.** Antibody Transfer Ratios and Cord Blood and Maternal Sera Levels at Delivery for Primary 2-Dose and Booster Groups.

| Lab Assay | Group | Transfer Ratio<br>Median (IQR) | Cord Blood Serum<br>GMT<br>(95% CI) | Maternal Serum<br>GMT<br>(95% CI) |
| --- | --- | --- | --- | --- |
| Spike IgG BAU/mL | Primary | 1.77 (1.38, 2.25) | 500.5<br>(424.1, 590.7) | 313.8<br>(260.7, 377.8) |
|  | Booster | 1.55 (1.17, 1.98) | 3290.4<br>(2713.6, 3989.9) | 2200.6<br>(1764.9, 2743.8) |
| RBD IgG BAU/mL | Primary | 1.76 (1.33, 2.15) | 673.1<br>(564.5, 802.6) | 428.6<br>(351.9, 522.1) |
|  | Booster | 1.58 (1.05, 1.93) | 5005.7<br>(4126.5, 6072.1) | 3382.9<br>(2703.7, 4232.86) |
| D614G nAb ID50* | Primary | 1.10 (0.76, 1.89) | 96.4<br>(80.2, 115.9) | 83.4<br>(65.4, 106.4) |
|  | Booster | 1.78 (1.12, 2.25) | 742.5<br>(588.5, 936.8) | 446.4<br>(342.1, 582.5) |
| Delta nAb ID50* | Primary | 1.10 (1.00, 1.69) | 52.1<br>(42.3, 64.2) | 39.0<br>(30.9, 49.2) |
|  | Booster | 1.74 (1.06, 2.38) | 644.5<br>(507.5, 818.4) | 390.2<br>(299.2, 508.9) |
| Omicron nAb ID50* | Primary | 1.00 (1.00, 1.00) | 13.8<br>(12.2, 15.6) | 13.5<br>(11.9, 15.3) |
|  | Booster | 1.69 (1.00, 2.75) | 109.2<br>(81.6, 146.1) | 60.2<br>(43.6, 83.1) |
| Pseudotyped WT nAb<br>ID50* | Primary | 2.36 (1.96, 2.84) | 220.8<br>(183.5, 265.7) | 93.4<br>(77.3, 112.9) |
|  | Booster | 1.79 (1.46, 2.20) | 941.4<br>(735.7, 1204.7) | 512.5<br>(399.1, 658.3) |

\*All data are included in the calculation of the transfer ratio including those below the LLOQ of the assay.

## References

1. Allotey J, Stallings E, Bonet M, et al. Clinical manifestations, risk factors, and maternal and perinatal outcomes of coronavirus disease 2019 in pregnancy: living systematic review and meta-analysis. BMJ. 2020 Sep 1;370:m3320. doi: 10.1136/bmj.m3320. PMID: 32873575; PMCID: PMC7459193.

2. Zambrano LD, Ellington S, Strid P, et al. Update: Characteristics of Symptomatic Women of Reproductive Age with Laboratory-Confirmed SARS-CoV-2 Infection by Pregnancy Status — United States, January 22–October 3, 2020. MMWR Morb Mortal Wkly Rep 2020;69:1641–1647. DOI: http://dx.doi.org/10.15585/mmwr.mm6944e3.

3. Marks KJ, Whitaker M, Agathis NT, et al. Hospitalization of Infants and Children Aged 0-4 Years with Laboratory-Confirmed COVID-19-COVID-NET, 14 States, March 2020-February 2022. MMWR Morb Mortal Wkly Rep 2022;71:429–436. doi: http://dx.doi.org/10.15585/mmwr.mm7111e2.

4. Carlsen EØ, Magnus MC, Oakley L, et al. Association of COVID-19 Vaccination During Pregnancy With Incidence of SARS-CoV-2 Infection in Infants. JAMA Intern Med. 2022 Jun 1. doi: 10.1001/jamainternmed.2022.2442. Epub ahead of print. PMID: 35648413.

5. The American College of Obstetricians and Gynecologists (ACOG). COVID-19 Vaccination Considerations for Obstetric-Gynecologic Care [Internet]. December 2020. Last Updated April 28, 2020. Available from: https://www.acog.org/clinical/clinical-guidance/practice-advisory/articles/2020/12/covid-19-vaccination-considerations-for-obstetric-gynecologic-care.

6. Munoz FM, Beigi RH, Posavad CM, et al. Multi-site observational maternal and infant COVID-19 vaccine study (MOMI-vax): a study protocol. BMC Pregnancy Childbirth. 2022 May 12;22(1):402. doi: 10.1186/s12884-022-04500-w. PMID: 35550037; PMCID: PMC9096328.

7. Gilbert PB, Montefiori DC, McDermott AB, et al. Immune correlates analysis of the mRNA-1273 COVID-19 vaccine efficacy clinical trial. Science. 2022 Jan 7;375(6576):43–50. doi: 10.1126/science.abm3425. Epub 2021 Nov 23. PMID: 34812653; PMCID: PMC9017870.

8. Edara VV, Pinsky BA, Suthar MS, et al. Infection and Vaccine-Induced Neutralizing-Antibody Responses to the SARS-CoV-2 B.1.617 Variants. N Engl J Med. Aug 12 2021;385(7):664–666. doi:10.1056/NEJMc2107799.

9. Belik M, Jalkanen P, Lundberg R, et al. Comparative analysis of COVID-19 vaccine responses and third booster dose-induced neutralizing antibodies against Delta and Omicron variants. Nat Commun. 2022 May 5;13(1):2476. doi: 10.1038/s41467-022-30162-5. PMID: 35513437; PMCID: PMC9072399.

10. Omer SB. Maternal Immunization. N Engl J Med. 2017 Mar 30;376(13):1256–1267. doi: 10.1056/NEJMra1509044. PMID: 28355514.

11. Centers for Disease Control (CDC). Interim Clinical Considerations for Use of COVID-19 Vaccines Currently Approved or Authorized in the United States. Last reviewed on May 23, 2022. Available from: https://www.cdc.gov/vaccines/covid-19/clinical-considerations/interim-considerations-us.html.

## References

1. Gilbert PB, Montefiori DC, McDermott AB et al. Immune correlates analysis of the mRNA-1273 COVID-19 vaccine efficacy clinical trial. Science. 2022 Jan 7;375(6576):43–50. doi: 10.1126/science.abm3425. Epub 2021 Nov 23. PMID: 34812653; PMCID: PMC9017870.

2. Alter G, Yu J, Liu J et al. Immunogenicity of Ad26.COV2.S vaccine against SARS-CoV-2 variants in humans. Nature. 2021 Aug;596(7871):268–272. doi: 10.1038/s41586-021-03681-2. Epub 2021 Jun 9. PMID: 34107529; PMCID: PMC8357629.

3. Pegu A, O’Connell SE, Schmidt SD et al. Durability of mRNA-1273 vaccine-induced antibodies against SARS-CoV-2 variants. Science. 2021 Sep 17;373(6561):1372–1377. doi: 10.1126/science.abj4176. Epub 2021 Aug 13. PMID: 34385356; PMCID: PMC8691522.

4. Self WH, Tenforde MW, Rhoads JP, et al. Comparative Effectiveness of Moderna, Pfizer-BioNTech, and Janssen (Johnson & Johnson) Vaccines in Preventing COVID-19 Hospitalizations Among Adults Without Immunocompromising Conditions — United States, March–August 2021. MMWR Morb Mortal Wkly Rep 2021;70:1337–1343. DOI: http://dx.doi.org/10.15585/mmwr.mm7038e1.

5. Demonbreun AR, Sancilio A, Velez ME et al. COVID-19 mRNA Vaccination Generates Greater Immunoglobulin G Levels in Women Compared to Men. J Infect Dis. 2021 Sep 1;224(5):793–797. doi: 10.1093/infdis/jiab314. PMID: 34117873; PMCID: PMC8536925.

6. Edara VV, Norwood C, Floyd K et al. Infection-and vaccine-induced antibody binding and neutralization of the B.1.351 SARS-CoV-2 variant. Cell Host Microbe. 2021 Apr 14;29(4):516–521.e3. doi: 10.1016/j.chom.2021.03.009. Epub 2021 Mar 20. PMID: 33798491; PMCID: PMC7980225.

7. Bajema KL, Dahl RM, Evener SL, et al. Comparative Effectiveness and Antibody Responses to Moderna and Pfizer-BioNTech COVID-19 Vaccines among Hospitalized Veterans — Five Veterans Affairs Medical Centers, United States, February 1–September 30, 2021. MMWR Morb Mortal Wkly Rep 2021;70:1700–1705. DOI: http://dx.doi.org/10.15585/mmwr.mm7049a2.

8. Fluss R, Faraggi D, Reiser B. Estimation of the Youden Index and its associated cutoff point. Biom J. 2005 Aug;47(4):458–72. doi: 10.1002/bimj.200410135. PMID: 16161804.

9. Cristiano A, Nuccetelli M, Pieri M et al. Serological anti-SARS-CoV-2 neutralizing antibodies association to live virus neutralizing test titers in COVID-19 paucisymptomatic/symptomatic patients and vaccinated subjects. Int Immunopharmacol. 2021 Dec;101(Pt B):108215. doi: 10.1016/j.intimp.2021.108215. Epub 2021 Oct 4. PMID: 34649115; PMCID: PMC8487771.

10. Jakuszko K, Kościelska-Kasprzak K, Żabińska M et al. Immune Response to Vaccination against COVID-19 in Breastfeeding Health Workers. Vaccines (Basel). 2021 Jun 17;9(6):663. doi: 10.3390/vaccines9060663. PMID: 34204501; PMCID: PMC8235492.

11. Edara VV, Pinsky BA, Suthar MS et al. Infection and Vaccine-Induced Neutralizing-Antibody Responses to the SARS-CoV-2 B.1.617 Variants. N Engl J Med. 2021 Aug 12;385(7):664–666. doi: 10.1056/NEJMc2107799. Epub 2021 Jul 7. PMID: 34233096; PMCID: PMC8279090.

12. Edara VV, Hudson WH, Xie X, Ahmed R, Suthar MS. Neutralizing Antibodies Against SARS-CoV-2 Variants After Infection and Vaccination. JAMA. 2021 May 11;325(18):1896–1898. doi: 10.1001/jama.2021.4388. PMID: 33739374; PMCID: PMC7980146.

13. Vanderheiden A, Edara VV, Floyd K et al. Development of a Rapid Focus Reduction Neutralization Test Assay for Measuring SARS-CoV-2 Neutralizing Antibodies. Curr Protoc Immunol. 2020 Dec;131(1):e116. doi: 10.1002/cpim.116. PMID: 33215858; PMCID: PMC7864545.

14. Katzelnick LC, Coello Escoto A, McElvany BD et al. Viridot: An automated virus plaque (immunofocus) counter for the measurement of serological neutralizing responses with application to dengue virus. PLoS Negl Trop Dis. 2018 Oct 24;12(10):e0006862. doi: 10.1371/journal.pntd.0006862. PMID: 30356267; PMCID: PMC6226209.

